# Comparing the of prevalence of Sarcopenia in 60 + population using different diagnostic criteria in North Indian population of India

**DOI:** 10.1101/2025.05.31.25328663

**Authors:** Yathath Malik, Minakshi Dhar, Yogesh Bahurupi, Shalini Sharma, Monika Pathania

## Abstract

**Introduction:** Sarcopenia refers to an age-related loss in muscular mass, strength, and function. Given global aging trends, the load is anticipated to increase. Prevalence estimates vary due to different diagnostic criteria by different organizations. This study uses AWGS 2019 criteria to calculate sarcopenia prevalence and compares it to FNIH, EWGSOP2, and SWAG-SARCO definitions to highlight differences in case identification and analyze their consequences for clinical management.

**Methodology:** This cross-sectional study was carried out among older population in Doiwala and Raipur Blocks in Dehradun district, Uttarakhand, India, using a multistage sampling method. Sarcopenia was analyzed using AWGS 2019 criteria and compared to the FNIH, EWGSOP2, and SWAG-SARCO definitions. Muscle mass, strength, and physical performance were measured using bioelectric impedance analysis (BIA), handgrip strength, and gait speed tests. After ethical approval and informed consent, structured interviews were used to obtain information about socio-demography, comorbidities, and lifestyle factors.

**Results:** 675 participants, with a mean age of 70.28 ± 7.88 years, mostly aged 60-69 were enrolled. Females accounted for 59.9% of the participants, most likely due to Uttarakhand’s hill-plain migration tendencies. Hypertension (36.3%) was the most prevalent co-morbidity. Cataracts were present in 13.3% of participants, and 54% were exposed to chulha smoke, the majority of whom were females. Using AWGS 2019, 23.37% were sarcopenic, with 6.51% having severe sarcopenia. EWGSOP2 diagnosed 18.22% of cases as likely sarcopenia, while FNIH diagnosed 5.18%. SWAG SARCO 2021 identified 17.63% of participants as sarcopenic. SWAG SARCO demonstrated greater diagnostic value over FNIH in the Indian context, with sensitivity and specificity of 61.33% and 94.86%, respectively. These data support SWAG SARCO as a valid method for detecting sarcopenia in community-dwelling elderly.

**Key Message:** Sarcopenia is prevalent in Uttarakhand’s older population and with changing diagnostic criteria producing different results, undermines the necessity for region-specific screening techniques.

## Introduction

Blooming from the gerontology research to clinical geriatric practice the term sarcopenia has gained traction with internists, gastroenterologists, pulmonologists and many more sub-specialties of medicine. Rosenberg coined the term ‘sarcopenia’ (1). Sarcopenia as a word has Greek origins. Sarcomeaning flesh and peniadeficiency, literally meaning the deficiency of flesh. With an ageing globe and human population, it is likely to become compounded to significantly higher proportions. Sarcopenia is an ailment marked by reduced muscle mass, strength, along with function. Various standards such as muscular mass, function, as well as strength in diverse combinations have defined sarcopenia. The Asian Working Group for Sarcopenia (AWGS) in 2019 has defined sarcopenia as “age related loss of muscle mass, plus low muscle strength, and/or low physical performance” (2). The EWGSOP 2 group prioritizes lower muscular strength compared to low muscle mass, indicating sarcopenia. Sarcopenia is defined by this group as the existence of insufficient muscular volume or function. Owing to the existence of reduced muscular strength, quantity/quality of muscles, along with poor physical performance defines severe sarcopenia (3). FNIH defines sarcopenia as a combination of reduced handgrip strength plus decreased muscle mass. The series analysis suggests following cut points for weakness: Grip strength <26kg for males and <16kg for females, also, adjusted for body mass index, appendicular lean mass <0.789 for males and <0.512 for females (4). Sarcopenia, according to SWAG-SARCO 2021, is a syndrome in which either of the listed criteria are unacceptable, as measured by muscular function, strength, and mass are all assessed using clinical, biochemical, and/or imaging techniques. (5). Sarcopenia prevalence varies widely because different research utilizes different criteria and cutoffs for assessing sarcopenia.

This study aimed to determine the prevalence of sarcopenia based on the AWGS 2019 consensus statement and later compare it to the prevalence defined by the Foundation for the National Institutes of Health (FNIH), the revised European Working Group on Sarcopenia in Older People (EWGSOP2), and the South Asian Working Action Group on Sarcopenia (SWAG-SARCO) criteria.

## Methods

### Study design

This cross-sectional study was conducted among the older population residing in Doiwala and Raipur Blocks in Dehradun district, Uttarakhand, India. Drawing on the Kadoma Sarcopenia study by Kurose S. et al. (6), prevalence of sarcopenia is 22.3%, assuming a confidence interval of 95%, Z value of 1.96 and relative precision (d) of 4.46%, using formula n= Z2 P(1-P)/ d 2 we calculated a sample size of 353 participants for our study, but we studied a cohort of 712 participants as identified under previous study conducted in the department. Out of these 712, 675 were enrolled in the study, as 12 had migrated to different places, 14 had died and remaining 11 had a SARC F < 4. We included community dwelling people of age more than or equal to 60 years and excluded terminally ill and bed ridden patients. The cutoff’s as per AWGS 2019, EWGSOP2, FNIH and SWAG SARCO criteria that were used for comparing prevalence of sarcopenia as shown in Table 1 (supplementary file).

**Table 1:** Sensitivity along with Specificity of EWGSOP 2 criteria for diagnosing Sarcopenia.

| Statistics | Value | 95% CI |
| --- | --- | --- |
| Sensitivity | 82.00% | 74.90% to 87.79% |
| Specificity | 100.00% | 99.30% to 100.00% |
| Positive Likelihood Ratio | — | — |
| Negative Likelihood Ratio | 0.18 | 0.13 to 0.25 |
| Disease prevalence (*) | 22.22% | 19.14% to 25.55% |
| Positive Predictive Value (*) | 100.00% | 97.05% to 100.00% |
| Negative Predictive Value (*) | 95.11% | 93.25% to 96.47% |
| Accuracy (*) | 96.00% | 94.23% to 97.35% |

Before beginning the study procedure, all patients were given signed informed consent. The study followed ethical principles in conformity with the aforementioned Declaration of Helsinki, Good Clinical Practice guidelines, along with all relevant local laws and regulations. It had been ethically cleared by the Institutional Ethics Committee under letter no. AIIMS/IEC/23/245 dated 19.07.2022.

## Procedure

### Sarcopenia screening

SARC-F had been used to screen sarcopenia. Five characteristics have been evaluated by the SARC-F questionnaire: strength, walking assistance, chair raising, stair climbing, and falls. Every component is graded from 0 to 2, with a minimum of 0 and a maximum of 10. Those with score of >4 is considered positive for sarcopenia screening (7).

### Muscle strength assessment

A “Jamar Handheld Dynamometer” was utilized to test the dominant hand’s handgrip strength, plus mean of these two values was used to choose the subgroup that would be measured for the muscle mass. The AWGS 2019 criteria set diagnostic cutoffs for low muscle strength as follows: Handgrip weight: < 28.0 kg in men and < 18.0 kg in women (2).

### Assessment of muscle mass

It was calculated with the Bio-impedance Analyzer HBF-702T (Omeron, India). The approach indicated in Figure 6 (supplementary file) was followed exactly as described in the manufacturer’s instructions. “OMRON HEALTHCARE Co., Ltd. 53, Kunotsubo, Terado-cho, Muko, Kyoto, 617-0002 Japan” (8).

### Data Collection

A pre-formed Performa was used to collect socio-demographic details of the patient.

### Statistical analysis

MS Excel and SPSS version 23 were used to compile the study data. Mean was used as measure of central tendency for BMI, ASMI, SARC-F scores, Hand grip, weight and FM%. A descriptive analysis was done for elaborating the baseline characteristics of the cohort ranging from age, sex, residence, co-morbidity and exposures/addictions. The prevalence of sarcopenia was identified by calculating the ratio of affected patients to the total number of participants diagnosed with the relevant condition by the total number of study participants (n=675). FNIH and SWAG SARCO consensus definitions were compared with AWGS 2019 criteria for sarcopenia using 2×2 table to calculate their respective sensitivity, specificity, positive and negative predictive value in our cohort.

## Results

The mean age of the participants was 70.28 ± 7.88 years, with most of them occurring between the ages of 60 and 69. Notably, 13.78% and 1.78% of the participants were octogenarians and nonagenarians respectively. 88 participants were oldest old as per the WHO definition. A relatively bell-shaped distribution for age in our cohort was observed. Of the 675 participants, 59.9% (n=404) were females and 40.1% were males (n=271) as shown in the table 4 and figure 13. Possibly the increasingly prevalent hill- plain migration in the state of Uttarakhand contributed to the higher number of female participants encountered in the doorto-door survey conducted in the study. With male members, either travelling, migrating or actively working their respective occupations at the time of field visits. 33.6% (n=227) residing in the hilly terrains of Thano and 66.4% (n=448) residing in the relatively plain area of Raiwala. The hill and plains were considered as one the parameters while evaluating the associated factors for sarcopenia and sarcopenic obesity in the present cohort.

Systemic hypertension was the most common co-morbidity present in 36.3% participants (n=245). The current cohort had 59 and 108 subjects with coronary artery disease plus Type 2 diabetes mellitus respectively, amounting to 8.7% and 16% respectively in the present cohort. Cataract was the third leading co-morbidity affecting 13.3% (n=90) participants. Amongst the addictions and exposures, most common was chulha exposure in 54% of participants predominantly female participants due to cultural reasons. 27% participants were smokers in the current cohort.

### Diagnosis of Sarcopenia as per AWGS 2019 Algorithm

According to the AWGS 2019 criteria (Figure 1, supplementary figgure), probable sarcopenia was identified in 23.40% (n=158). The overall prevalence of sarcopenia was 22.37% (n=151), including 6.51% (n=44) with severe sarcopenia. The remaining 77.62% (n=524) were classified as non-sarcopenic.

**Figure 1:**
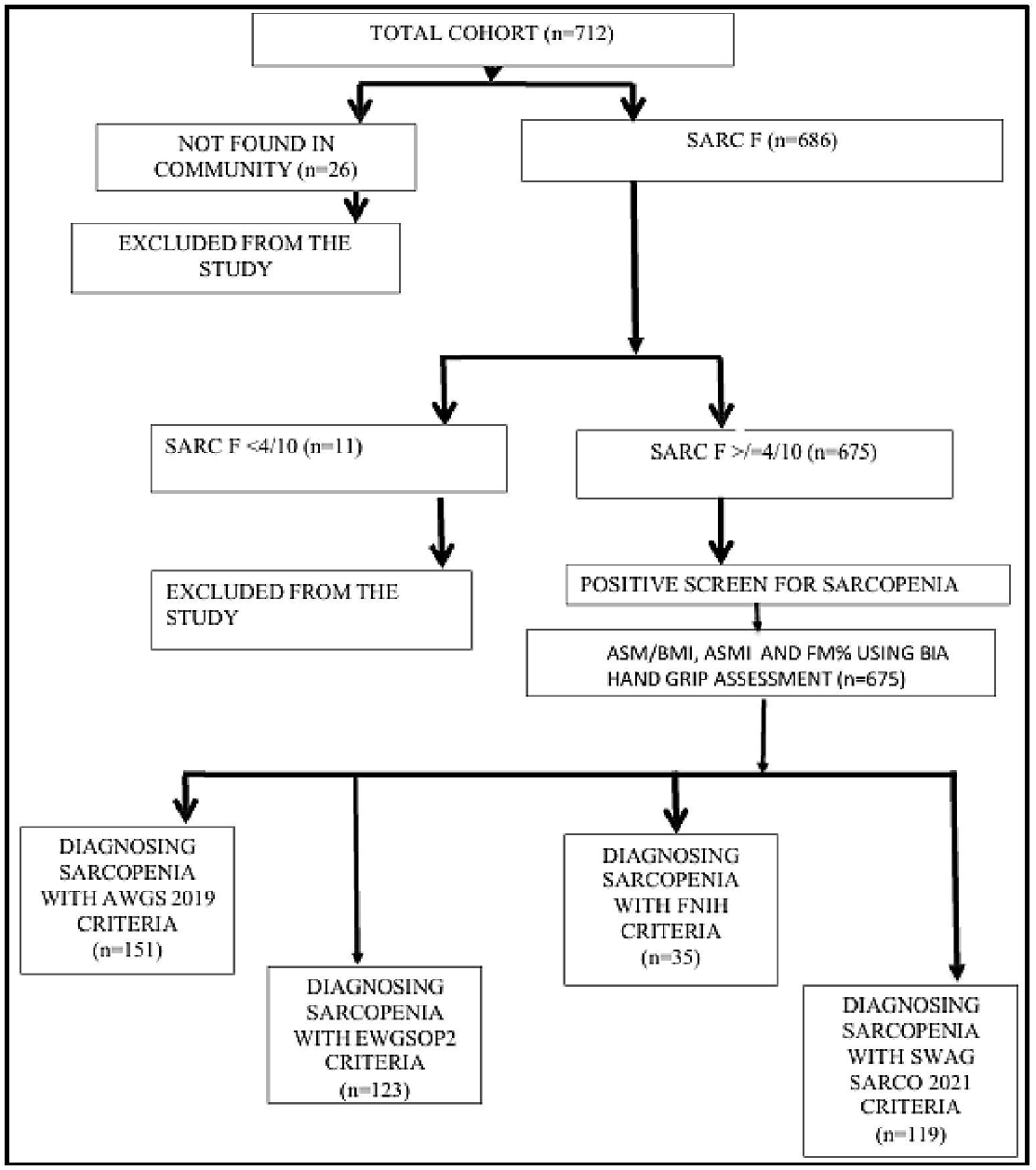
Studyflow in the present study.

### Diagnostic algorithm as per EWGSOP2 criteria

According to the EWGSOP2 consensus criteria (Figure 2, supplementary file), possible sarcopenia was identified in 17.92% (n=121). The prevalence of sarcopenia was 4.29% (n=29), with 2.37% (n=16) classified as having severe sarcopenia. The remaining 81.77% (n=552) were non-sarcopenic.

### Diagnostic algorithm as per FNIH criteria

According to the FNIH criteria (Figure 3, supplementary file), possible sarcopenia was identified in 16.44% (n=111), with a sarcopenia prevalence of 5.18% (n=35). The remaining 94.81% (n=640) were classified as non-sarcopenic.

### Diagnostic algorithm as per SWAG SARCO 2021

According to the SWAG-SARCO 2021 criteria (Figure 4, supplementary file), the overall prevalence of sarcopenia was 17.62% (n=119). Possible sarcopenia was observed in 23.40% (n=158), while 13.77% (n=93) had confirmed sarcopenia. Additionally, 5.02% (n=26) of cases were missed as possible sarcopenia. The remaining 76.59% (n=517) were classified as non-sarcopenic.

#### Diagnostic value of different criteria

Next, an attempt was made to shed some light on the diagnostic value of the various criteria/consensus for sarcopenia in our cohort. The AWGS 2019 criteria was selected as the gold standard as it has been validated in numerous studies for its applicability to the Asian population including India.

Table 1 presents the sensitivity alongside specificity of EWGSOP 2 as compared to AWGS 2019 criteria in diagnosing probable Sarcopenia.

Table 2 displays both specificity and sensitivity of the EWGSOP2 sarcopenia diagnostic criteria. Sensitivity and specificity came 19.21% and 100%, respectively. Regarding suspected sarcopenia, the EWGSOP2 is more sensitive and accurate than for sarcopenia. It likely stems from the different cut-offs for the involved parameters. As such, it would not be a good tool for screening sarcopenia in the Indian setting.

**Table 2:**
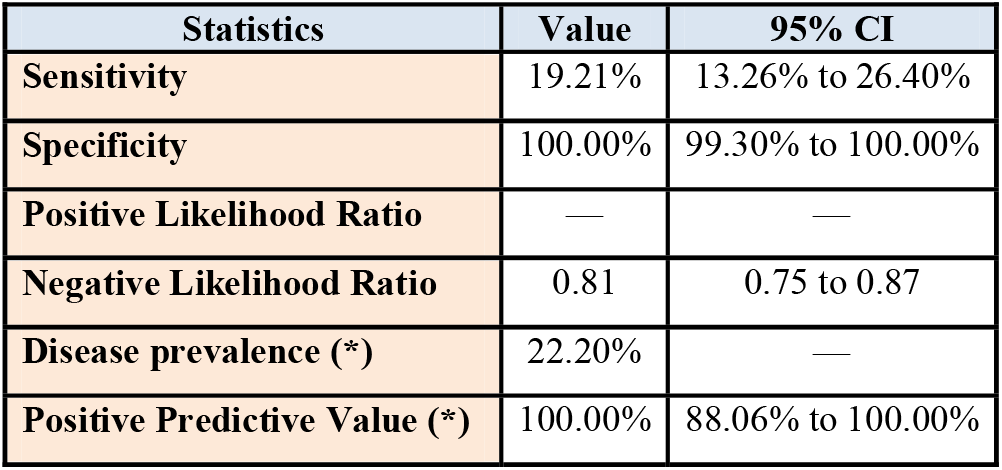
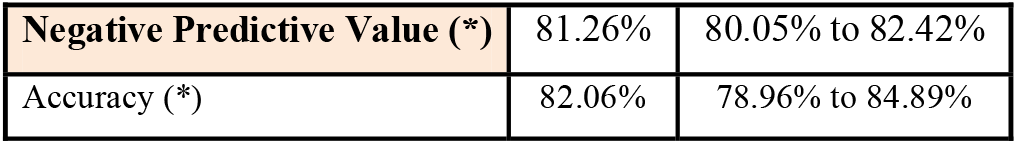
Specificity and Sensitivity of the EWGSOP2 criteria for diagnosing Sarcopenia.

| Statistics | Value | 95% CI |
| --- | --- | --- |
| Sensitivity | 19.21% | 13.26% to 26.40% |
| Specificity | 100.00% | 99.30% to 100.00% |
| Positive Likelihood Ratio | — | — |
| Negative Likelihood Ratio | 0.81 | 0.75 to 0.87 |
| Disease prevalence (*) | 22.20% | — |
| Positive Predictive Value (*) | 100.00% | 88.06% to 100.00% |
| **Negative Predictive Value (\*)** | 81.26% | 80.05% to 82.42% |
| **Accuracy (\*)** | 82.06% | 78.96% to 84.89% |

Table 3 displays both the sensitivity and specificity of the FNIH criteria, which were 23.3% and 100%, respectively. in diagnosing sarcopenia in our cohort with 95% confidence intervals of 16.82% to 30.93% and 99.30% to 100.00% respectively. The FNIH consensus seems unlikely to be a sensitive technique for screening sarcopenia in our group of community-dwelling seniors. However, true to its algorithm and definition, it has a 96% accuracy in diagnosing sarcopenia.

**Table 3:** Sensitivity and Specificity of FNIH criteria for diagnosis of Sarcopenia.

| **Statistics** | **Value** | **95% CI** |
| --- | --- | --- |
| **Sensitivity** | 23.33% | 16.82% to 30.93% |
| **Specificity** | 100.00% | 99.30% to 100.00% |
| **Positive Likelihood Ratio** | - |  |
| **Negative Likelihood Ratio** | 0.77 | 0.70 to 0.84 |
| **Positive Predictive Value** | 100.00% | 90.00% to 100.00% |
| **Negative Predictive Value** | 82.03% | 80.69% to 83.30% |
| **Accuracy** | 82.96% | 79.91% to 85.72% |

Regarding the diagnostic value of the SWAG-SARCO consensus, as presented in Table 5, it demonstrated a sensitivity of 61.33% (95% CI: 53.05%–69.16%) and a specificity of 94.86% (95% CI: 92.61%–96.58%) for diagnosing sarcopenia in comparison to ASWG 2019 criteria. It has a higher negative likelihood ratio of 0.41 than the ASWG 2019 criteria as shown in Table 4. The SWAG SARCO consensus is a better tool than FNIH to screen for sarcopenia with a higher sensitivity and is a more specific consensus than FNIH group for diagnosing sarcopenia in our representative population.

**Table 4:**
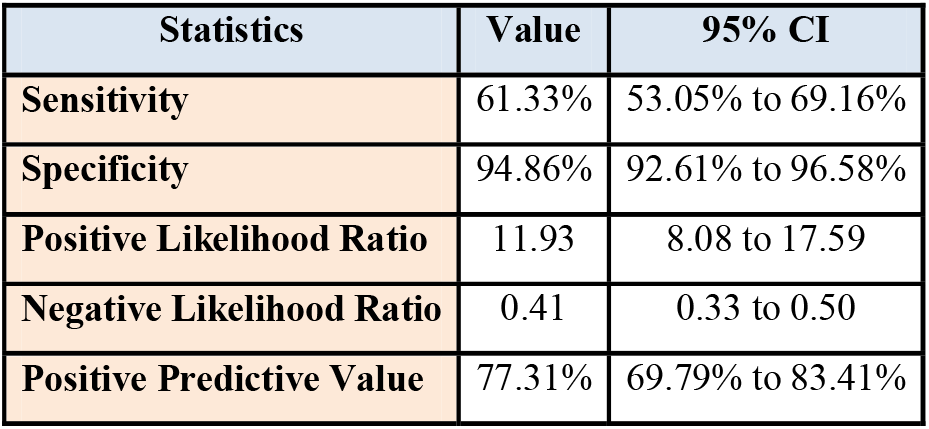
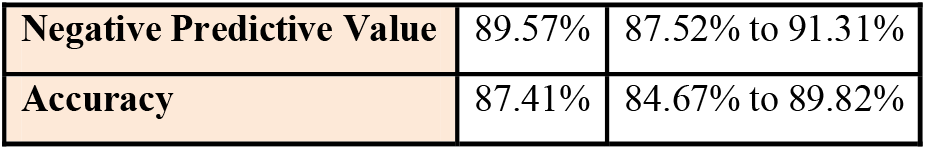
Sensitivity and Specificity of SWAG-SARCO criteria for diagnosis of Sarcopenia.

| **Statistics** | **Value** | **95% CI** |
| --- | --- | --- |
| **Sensitivity** | 61.33% | 53.05% to 69.16% |
| **Specificity** | 94.86% | 92.61% to 96.58% |
| **Positive Likelihood Ratio** | 11.93 | 8.08 to 17.59 |
| **Negative Likelihood Ratio** | 0.41 | 0.33 to 0.50 |
| **Positive Predictive Value** | 77.31% | 69.79% to 83.41% |
| **Negative Predictive Value** | 89.57% | 87.52% to 91.31% |
| **Accuracy** | 87.41% | 84.67% to 89.82% |

Epidemiologically, the diagnostic performance of SWAG SARCO and AWGS 2019 criteria/consensus are comparable for our cohort. Given the inherent upside of SWAG SARCO consensus in defining sarcopenia as a syndrome, with higher theoretical applicability in the South Asian and by extension Indian population, SWAG SARCO is a robust tool in diagnosing sarcopenia in the Indian context as shown in Table 4.

## Discussion

Sarcopenia is an intriguing geriatric syndrome, with a multi-pronged pathophysiology, multi-factorial etiology, multifaceted clinical presentation as well as multi-caveated treatment approach. This study noted a prevalence of 22.3% of probable sarcopenia as per the AWGS2019 criteria across the community residing elderly in the block areas of Thano and Raiwala in Uttarakhand. Translating to at least 1 in 5 elderly having probable sarcopenia. It is similar to the conclusions drawn by the Kadoma sarcopenia study from Korean elderly. They also reported 22.3% prevalence using the AWGS 2019 criteria in their retrospective analysis (6).

Ekasame Vanitcharoenkul1 et al. discovered that sarcopenia had an overall prevalence of 18.1%, with almost two-thirds (66.9%) of the 2,456 Thai participants classified as having severe sarcopenia based on the AWGS 2019 criteria. A large sample size, stratified multistage sampling method could have contributed to a lower overall prevalence (9).

In a Taiwanese study by Yu-Shiue Chen et al, 100 healthcare professionals with mean age of 41.8 ± 13.3 years, out of which 53% were females, reported a 22% prevalence of sarcopenia and 19% with presarcopenia. This could represent the natural progression from presarcopenia to sarcopenia in this young cohort. (10) However, a higher prevalence of 32% was reported by Lan-Anh Thi Pham from Vietnam using the AWGS 2019 cutoffs. Probably the different BIA device, genetic and socioeconomic differences could have contributed to this. (11) However, closer home, Altaf et al, from Pakistan reported a prevalence of sarcopenia to be 47.18% among 142 elderly recruited in Islamabad. The design differences- DEXA scan to calculate SMI, not using performance and muscle strength parameters, definition of cut-offs other than the AWGS 2019 criteria could have contributed to this high prevalence. (12)

Charmaine Tan You Mei et al. identified 383 patients who were hospitalized in post-acute hospital in Singapore. Sarcopenia was prevalent in 54% of cases, with severe sarcopenia accounting for 38.9%. The acute illness could have weakened the muscular strength as well as the physical performance indices, resulting a higher prevalence in this population with acute on chronic sarcopenia (13). In Egypt, 69.7% of elderly men and postmenopausal women over 50 admitted with osteoporotic fractures (hip fractures or severe osteoporosis) were diagnosed with sarcopenia using the Arabic SARC-F score. The inherent differences in the design, i.e. including a population with multiple risk factors of secondary sarcopenia and using Arabic SARC F tool could have contributed to these stark differences (14).

From India, 678 elderly patients coming as out-patients to the department of medicine at Kalinga Institute of Medicine in Odisha has a prevalence of 15.3% among old males and 20.5% among females. They had a higher number of elderly males than the current study and no defined cut-offs as published by the AWGS, SWAG SARCO or other groups were used. These could explain the lower prevalence as reported by Mohanty L et al in 2016 (15). According to the LASI, 936 people (3.4%) exhibited both sarcopenia and diabetes, while 6412 (23.5%) reported only sarcopenia., 3190 (11.7%) had only diabetes, and the vast majority, 16,703 (61.3%), had neither. Sarcopenia was found in 7348 people, giving a population frequency of 27.0%. A bigger, more diverse sample size could have contributed to a larger population prevalence for sarcopenia than the current study. However, 23.5% individuals who had sarcopenia probably is the true estimate, because the remaining have diabetes along with sarcopenia, leading to an inflated population prevalence. The proportion of diabetics in the current study is lower than one published under LASI, explaining the differences in the prevalence between the 2 cohorts (16). As previously reported, the myriads of definitions proposed by various groups and the use of varying cut-offs contributed to the wide range of sarcopenia prevalence reported in the literature. Overall, this study resonates with previous literature that around 1 in 5 community dwelling elderly have probable sarcopenia. This brings light to the fact that screening for sarcopenia among the older adults at the grassroots involving the subcenters, primary and community health centers across the country should aid in early diagnosis, treatment and comprehensive rehabilitation of the ageing Indians.

To the best of the authors’ knowledge, this study stands among the pioneering efforts to explore different consensual definitions of sarcopenia among elderly population in the communities of India, as demonstrated in the results section. Asian Working Group for Sarcopenia 2019 criterion, combined with SWAG SARCO consensus criterion are most appropriate for diagnosing sarcopenia in the current cohort’s representative population. EWGSOP2 is a specific criterion for diagnosing both probable sarcopenia and sarcopenia, however has a lower sensitivity. However, FNIH clinical algorithm is not a robust tool for defining sarcopenia in elderly population residing in the community.

The authors agree that well designed cohort studies including population at risk of primary sarcopenia would dwell light on the true prevalence of these geriatric syndromes. As aforementioned, our study is in fair agreement with other Indian studies on the topic.

The authors also agree with other groups in the fact that the patho-biology and hence the formal objective definitions need a thorough re-look in the purview of this important geriatric syndrome. But at the same time, regional, socio-political, economic and diet patterns which have a vast diversity in our country could hinder in direct comparisons with the foreign studies. Another aspect deems attention that these discrepancies might not be repressed, rather should be acknowledged by the medical fraternity. The authors understand that as with other geriatric syndromes of frailty, falls, cognitive impairment, inter-patient variability is seen in the clinics, we conclude that similar person to person, cohort to cohort, country to country and inter-continent variations should be embraced gracefully and regional operational definitions which have been validated for the specific population subgroup should be devised and implemented in routine geriatric practice and community alike till the time patho-biology of sarcopenia is fully elucidated by metabolic, genomic and epigenetic studies in the coming future.

## Supporting information

Splemental Figure 4

## Data Availability

Data will be available upon reasonable request

