## Supplementary figures and images for "Comparing the of prevalence of Sarcopenia in 60 + population using different diagnostic criteria in North Indian population of India"

### Figure 1 supple.docx

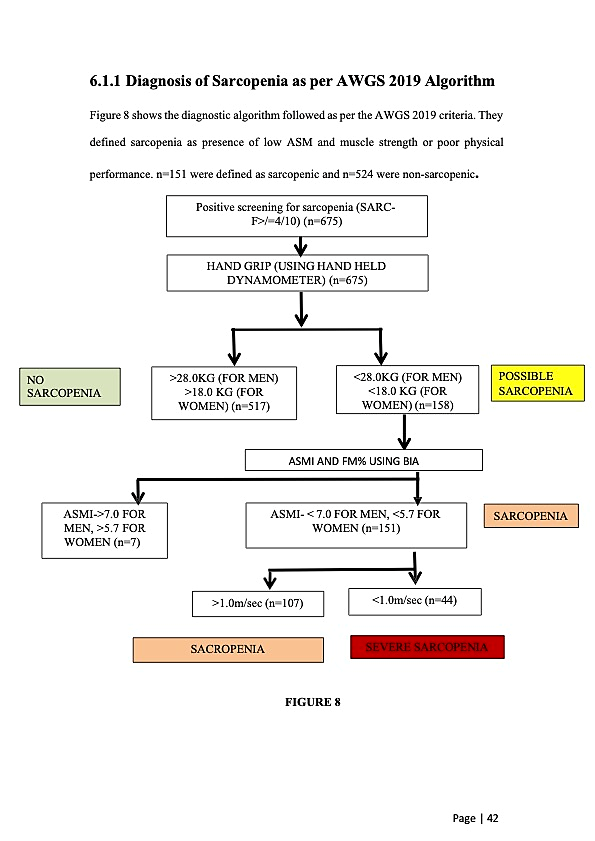

**Figure 1:Diagnosis of Sarcopenia as per AWGS 2019 Algorithm**

### Figure 2 supple.docx

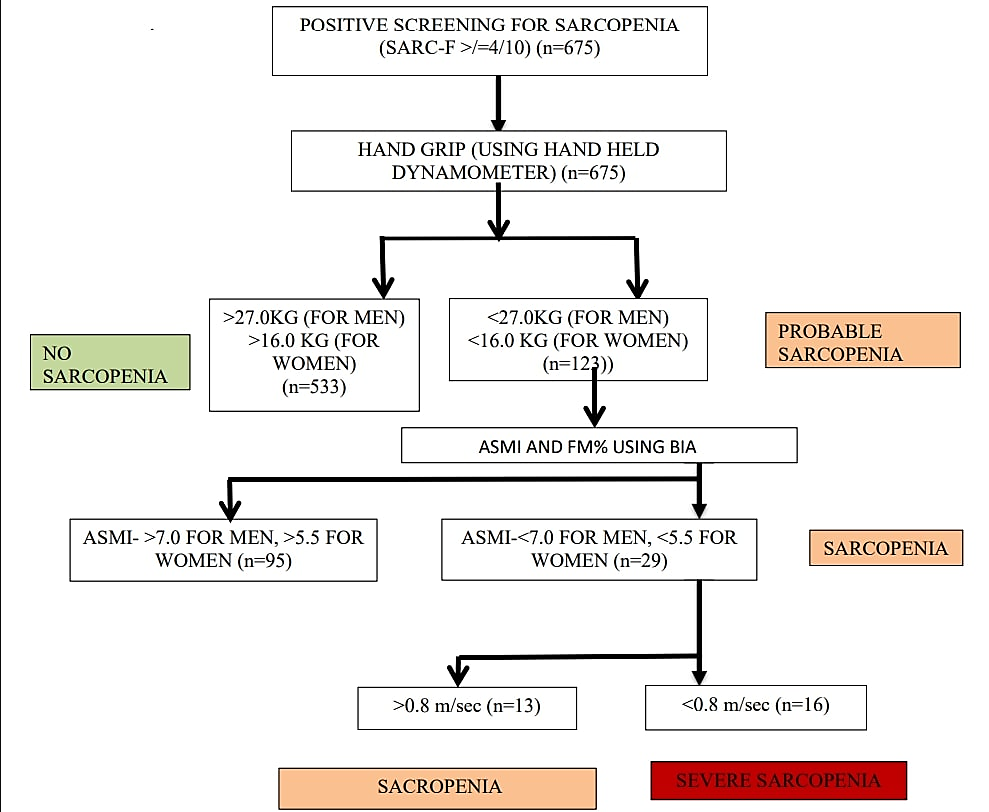

**Figure 2:Diagnostic algorithm as per EWGSOP2 criteria**

### Figure 3 supple.docx

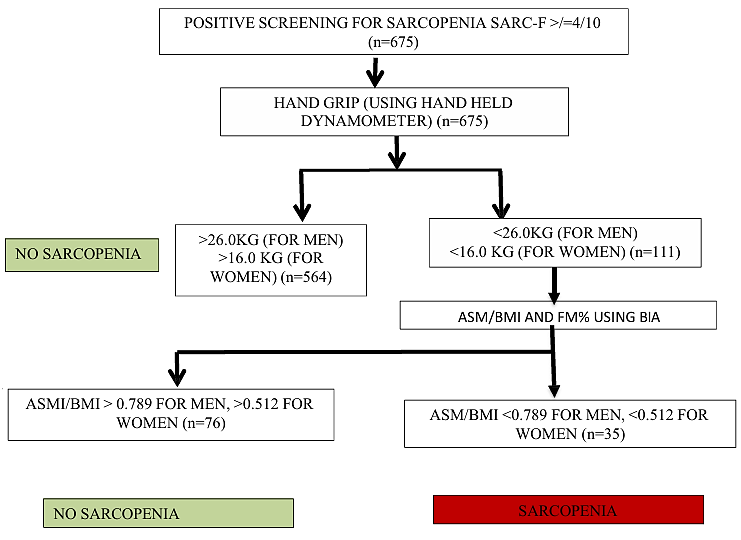

**Figure 3:Diagnostic algorithm as per FNIH criteria**

### Figure 4 supple.docx

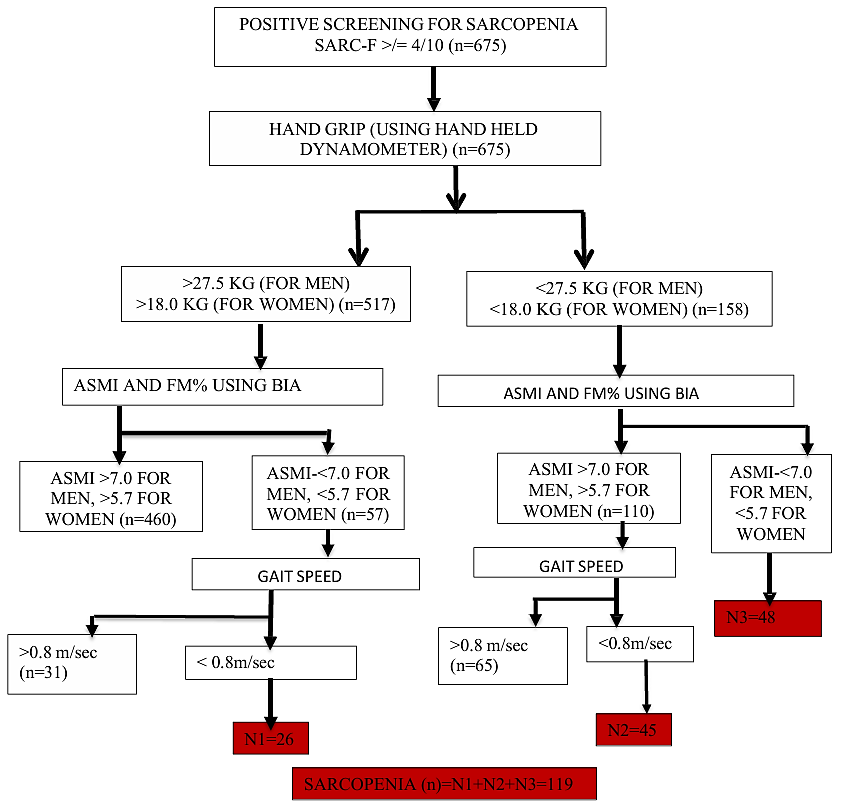

**Figure 4:Diagnostic algorithm as per SWAG SARCO 2021**
