## Supplementary material for "Comparing the of prevalence of Sarcopenia in 60 + population using different diagnostic criteria in North Indian population of India": Suplemental Table 1: Table 1 supple.docx

**Table 1:Comparative diagnostic criteria from various consensus statements**

| **Parameter** | **AWGS 2019** | | **SWAG SARCO** | | **EWGSOP2** | | **FNIH** | |
| --- | --- | --- | --- | --- | --- | --- | --- | --- |
|  | **Men** | **Women** | **Men** | **Women** | **Men** | **Women** | **Men** | **Women** |
| **SARC-F** | >4/10 | | >4/10 | | >4/10 | | NA | |
| **Hand grip** | <28 kg | <18 kg | <27.5 kg | <18 kg | <27 kg | <16 kg | <26 kg | <16 kg |
| **Gait speed** | <1.0 m/s | | <0.8 m/s | | <0.8 m/s | | NA | |
| **ASM** | NA | | NA | | NA | |  |  |
| **ASMI** |  |  |  |  | <20 kg | <15 kg |  |  |
| **ASM/BMI** | <7.0 kg/m² | <5.7 kg/m² | <7.0 kg/m² | <5.7 kg/m² | <7.0 kg/m² | <5.5 kg/m² | <0.789 | <0.512 |
